# The challenge of SARS-CoV-2 environmental monitoring in schools using floors and portable HEPA filtration units: Fresh or relic RNA?

**DOI:** 10.1101/2021.11.12.21266178

**Authors:** Rogelio Zuniga-Montanez, David A. Coil, Jonathan A. Eisen, Randi Pechacek, Roque G. Guerrero, Minji Kim, Karen Shapiro, Heather N. Bischel

## Abstract

Testing surfaces in school classrooms for the presence of SARS-CoV-2, the virus that causes COVID-19, can provide public-health information that complements clinical testing. We monitored the presence of SARS-CoV-2 RNA in five schools (96 classrooms) in Davis, California (USA) by collecting weekly surface-swab samples from classroom floors and/or portable high-efficiency particulate air (HEPA) units. Twenty-two surfaces tested positive, with qPCR cycle threshold (Ct) values ranging from 36.07–38.01. Intermittent repeated positives in a single room were observed for both floor and HEPA filter samples for up to 52 days, even following regular cleaning and HEPA filter replacement after a positive result. We compared the two environmental sampling strategies by testing one floor and two HEPA filter samples in 57 classrooms at Schools D and E. HEPA filter sampling yielded 3.02% and 0.41% positivity rates per filter sample collected for Schools D and E, respectively, while floor sampling yielded 0.48% and 0% positivity rates. Our results indicate that HEPA filter swabs are more sensitive than floor swabs at detecting SARS-CoV-2 RNA in interior spaces. During the study, all schools were offered weekly free COVID-19 clinical testing. On-site clinical testing was offered in Schools D and E, and upticks in testing participation were observed following a confirmed positive environmental sample. However, no confirmed COVID-19 cases were identified among students associated with classrooms yielding positive environmental samples. The positive samples detected in this study appeared to reflect relic viral RNA from individuals infected before the monitoring program started and/or RNA transported into classrooms via fomites. The high-Ct positive results from environmental swabs further suggest the absence of active infections. Additional research is needed to differentiate between fresh and relic SARS-CoV-2 RNA in environmental samples and to determine what types of results should trigger interventions.

## 1 Introduction

Early in the COVID-19 pandemic there were broad concerns that surfaces might serve as a source of transmission of SARS-CoV-2, the causal agent of the widespread infectious disease. However, while some early studies suggested that SARS-CoV-2 virus could remain infectious on surfaces for days (Kasloff *et al*., 2020; Riddell *et al*., 2020; van Doremalen *et al*., 2020; Harvey *et al*., 2021), there has been no indisputable evidence for surface-to-person transmission. Nevertheless, the stability of SARS-CoV-2 RNA on surfaces suggests that environmental monitoring via surface swabs could support the COVID-19 response.

Screening environmental samples such as wastewater and surface swabs for the presence of SARS-CoV-2 RNA provides indirect evidence of the number of infected people shedding the virus in the vicinity (Marshall *et al*., 2020; Betancourt *et al*., 2021; Renninger *et al*., 2021; Vo *et al*., 2021). Because reverse transcriptase quantitative polymerase chain reaction (RT-qPCR) tests for SARS-CoV-2 RNA are widely available and relatively inexpensive, environmental monitoring has become a cost-effective complement to clinical testing (Thompson *et al*., 2020; Harvey *et al*., 2021). Environmental monitoring also avoids issues associated with informed consent, sample collection, operational logistics, and equity that can slow or constrain clinical-testing programs (McElfish *et al*., 2021; Vandenberg *et al*., 2021).

The value of environmental monitoring has been most clearly demonstrated with wastewater surveillance (Medema *et al*., 2020; Wu *et al*., 2021). But while wastewater surveillance can inform policy and action at regional, city, neighborhood, and building levels, it cannot provide information about virus presence in interior spaces (e.g., building floors and rooms) and can provide only very limited assistance in identifying potential virus exposures. Surface sampling is a type of environmental monitoring that could fill this gap by providing decision-makers information at another level of resolution (between larger-scale environmental monitoring and individual clinical testing).

Environmental sampling for SARS-CoV-2 through high-touch surface testing has been examined by both academic research-based projects (Zhou *et al*., 2020; Harvey *et al*., 2021) as well as via companies offering monitoring services (Marshall *et al*., 2020). Several groups, including ours, have also evaluated viral RNA in HVAC systems with mixed results (Mouchtouri *et al*., 2020; Nissen *et al*., 2020; Coil *et al*., 2021; Horve *et al*., 2021; Maestre *et al*., 2021) or with portable air samplers (Ang *et al*., 2021). Complexities of HVAC systems (e.g., shared air between rooms, timed operation, difficult to access, variable filter types) have prevented filter-based monitoring from being widely deployed. Portable high-efficiency particulate air (HEPA) filtration units have the potential to be an effective mitigation strategy for SARS-CoV-2 transmission indoors (Curtius, Granzin and Schrod, 2021), and their widespread deployment through the pandemic creates a new opportunity for filter-based environmental monitoring.

In this study, we piloted a SARS-CoV-2 environmental monitoring study from January to August 2021 where we systematically collected floor and/or HEPA filter swab samples in five elementary schools. Swab samples were collected using oral swabs, and SARS-CoV-2 RNA was quantified through RT-qPCR. We compared the efficacy of floor and HEPA-filter samples for detecting SARS-CoV-2 RNA and COVID-19 cases in two of the schools. We hypothesized that HEPA filter sampling would be a more efficient strategy to detect infected individuals since SARS-CoV-2 virions would concentrate on the external surface of the filters as air circulated through the units.

## 2 Materials and Methods

### 2.1 Validation of SARS-CoV-2 detection on surfaces and HEPA air purification units

Prior to beginning the sampling campaign, we validated the detection of SARS-CoV-2 on surfaces using opportunistic sampling in two locations within six days after one or more clinical COVID-19 cases were identified. The sampling was conducted once in a house and twice in a school classroom. Forty-six samples were collected from different surfaces and tested, including a portable HEPA filtration unit (MA-40, Medify Air, USA) that was located in the classroom (Table S2). The HEPA filter was dismantled for testing, collecting samples from the outer grill cover, pre-filter mesh, and H13 HEPA filter surface. Detailed information on the validation sampling can be found in the supplementary information section S1.1.

### 2.2 Pilot study sampling framework and locations

We partnered with five schools in Davis, California, USA, to conduct weekly SARS-CoV-2 environmental monitoring using floor and HEPA filter swab samples from January to August 2021. All schools had in-person teaching and pandemic control plans and policies in place. One of two main sampling strategies was applied at each school; (1) only floor or (2) floor and HEPA filter sampling. The environmental sampling strategy at each school is summarized in Table 1. No personal information was collected on any individuals in the schools. The University of California, Davis IRB Administration determined that the study design was exempt from IRB review and approval.

**Table 1.** HEPA filter and floor environmental monitoring strategies for the detection of SARS-CoV-2 in five K-6 and K-8 schools.

| School (School type) | Preliminary testing period | Weekly sampling period | Concluding sampling episode | Number of classrooms sampled | Other rooms sampled | Enrollment | Samples collected per room and collection periods | Number of rooms with air purifiers (AP) sampled and AP models |
| --- | --- | --- | --- | --- | --- | --- | --- | --- |
| A (K-6) | - | January 28 to August 12, 2021 | - | 12 | 6 | 135-138 students | - One floor sample per room. January 28 to August 12, 2021<br>- One HEPA filter sample per room. May 20 to August 12, 2021 | - Rooms: 18<br>- AP models: 12 MA-40, Medify Air, USA. 6 AeraMax 300, Fellowes, USA |
| B (K-6) | - | January 28 to August 12, 2021 | - | 3 | 7 | 47-48 students | - One floor sample per room. January 28 to August 12, 2021<br>- One HEPA filter sample per room. May 20 to August 12, 2021 | - Rooms: 7<br>- AP models: MA-40, Medify Air, USA |
| C (K-8) | February 23, 2021 (11 rooms)<sup>a</sup> | March 2 to May 25, 2021 | - | 10 | 1 | 259 students | - One floor sample per room. March 2 to May 25, 2021 | - Rooms: 0 |
| D (K-6) | March 10 to March 30, 2021 (2 rooms)<sup>b</sup> | April 13 to June 08, 2021 | June 18, 2021 (8 rooms)<sup>d</sup> | 25 | - | 380 students | - One floor and two distinct HEPA filter samples per room. March 10 to June 18, 2021 | - Rooms: 25<br>- AP models: MA-40, Medify Air, USA |
| E (K-6) | March 10 to March 30, 2021 (1 room)<sup>c</sup> | April 13 to June 07, 2021 | - | 32 | - | 450 students | - One floor and two distinct HEPA filter samples per room. March 10 to June 07, 2021 | - Rooms: 32<br>- AP models: MA-40, Medify Air, USA |

<sup>a</sup>Preliminary testing conducted in the same rooms as weekly sampling; however, only a single surface per room was swabbed. These surfaces included walls, desks, sink counters, floors, door handles, cabinets, and tables.
<sup>b</sup>Preliminary testing conducted in Rooms 11 and 20.
<sup>c</sup>Preliminary testing conducted in a not included in the weekly sampling.
<sup>d</sup>Concluding sampling session at the end of the school year was conducted in Rooms 5, 6, 9, 13, 15, 16, 17 and 24. A door jam, teacher desk, center surface (desk or projector), floor, and two air filter samples were collected from each room.

### 2.3 HEPA filter and surfaces sampling

Environmental samples were collected using nylon fiber oral swabs with an ABS handle (Miraclean Technology Co. Ltd, China) that were pre-moistened in DNA/RNA Shield (Zymo Research, USA) before collecting the samples. For floor samples, a square area of approximately 10 cm × 10 cm in the center of the room was thoroughly sampled while rotating the swab. For other surfaces or items, a similar or smaller area, depending on surface or item size, was swabbed in a similar manner to floor samples. For the portable HEPA filters, the intake side filter cover was removed and the whole pre-filter mesh was thoroughly sampled by rotating the swab. After sample collection, the swab tip was snapped off by bending and rolling the swab at the 30 mm breakpoint without touching the sample. The swab tip was preserved in 500 μl of DNA/RNA Shield until laboratory processing. All surfaces sampled were wiped down with 75% ethanol wipes (Zhejiang Youquan Care Products Technology Co., Ltd., China) after sample collection.

The sampling at Schools A, B, C, D, and E was conducted by either the school or Healthy Davis Together (HDT) personnel and sampling kits were prepared and delivered to each location weekly. We created instructional videos to show the sampling teams how to collect HEPA filter and floor samples (supplementary information section S1.2).

### 2.4 RNA extraction and RT-qPCR for surface and air filter swab samples

Samples were received on the same day as sample collection, stored at room temperature for up to 4 hours and processed. DNA/RNA Shield transport media has been demonstrated to stabilize SARS-CoV-2 RNA at ambient temperatures for up to 28 days (FDA, 2020). Before RNA extraction, samples containing the swab tip in DNA/RNA Shield were vortexed at a medium-high to high speed for 10 minutes to suspend and homogenize the particles collected. The samples were then centrifuged at 10,000 x g for 1 minute to remove bubbles that formed during vortexing. Samples collected through February 11, 2021 were extracted manually utilizing the PureLink Viral RNA/DNA kit (Thermo Fisher Scientific, USA) according to manufacturer instructions and starting with a 200 μl sample volume. The validation test samples collected after known positive exposures, as well as all samples collected on March 30, 2021 were also manually extracted. Samples collected from February 12, 2021 through the end of the study were extracted using the MagMAX Microbiome Ultra Nucleic Acid Isolation Kit (Applied Biosystems, USA) and a KingFisher Flex automated purification system (Thermo Fisher Scientific, USA). The MagMAX_Microbiome_Stool_Flex.bdz nucleic acid isolation protocol (Applied Biosystems, USA) was utilized, with small modifications. In brief, the sample lysis step was not conducted as lysis was achieved through the use of DNA/RNA Shield and vortexing. The sample plate was loaded with 200 μl of sample and 260 μl of Binding Bead Mix. RNA extracts were eluted with 100 μl of Elution Solution and stored at -80 °C prior to RT-qPCR. The detailed manual and automated extraction protocols are available in the supplementary information section S1.3.

Extracts were thawed on ice after removal from -80 °C. All extracts were analyzed by RT-qPCR targeting the spike glycoprotein (S) gene of SARS-CoV-2 (Chan *et al*., 2020) using the Luna Universal Probe One-Step RT-qPCR Kit (New England Biolabs Inc., USA). Each 20 μl reaction contained 10 μl Luna Universal Probe One-Step Reaction Mix (2X), 1 μl Luna WarmStart RT Enzyme Mix (20X), specified concentrations of 1.5 μl combined primer/probe mix (Table 2), 2.5 μl nuclease-free water, and 5 μl RNA extract. Samples were analyzed in triplicate. Duplicates of positive (SARS-CoV-2 RNA extract donated by the University of Oregon) and no template (nuclease-free water) controls were included with each qPCR plate. The RT-qPCR assays were performed using the StepOnePlus Real-time PCR System (Applied Biosystems, USA). The thermal cycling conditions were 55 °C for 15 minutes and 95 °C for 2 minutes, followed by 40 cycles at 95 °C for 10 seconds and 60 °C for 60 seconds. Samples with at least one of three technical replicates with a cycle threshold (Ct) value lower than 40 were considered positive for SARS-CoV-2.

**Table 2.**
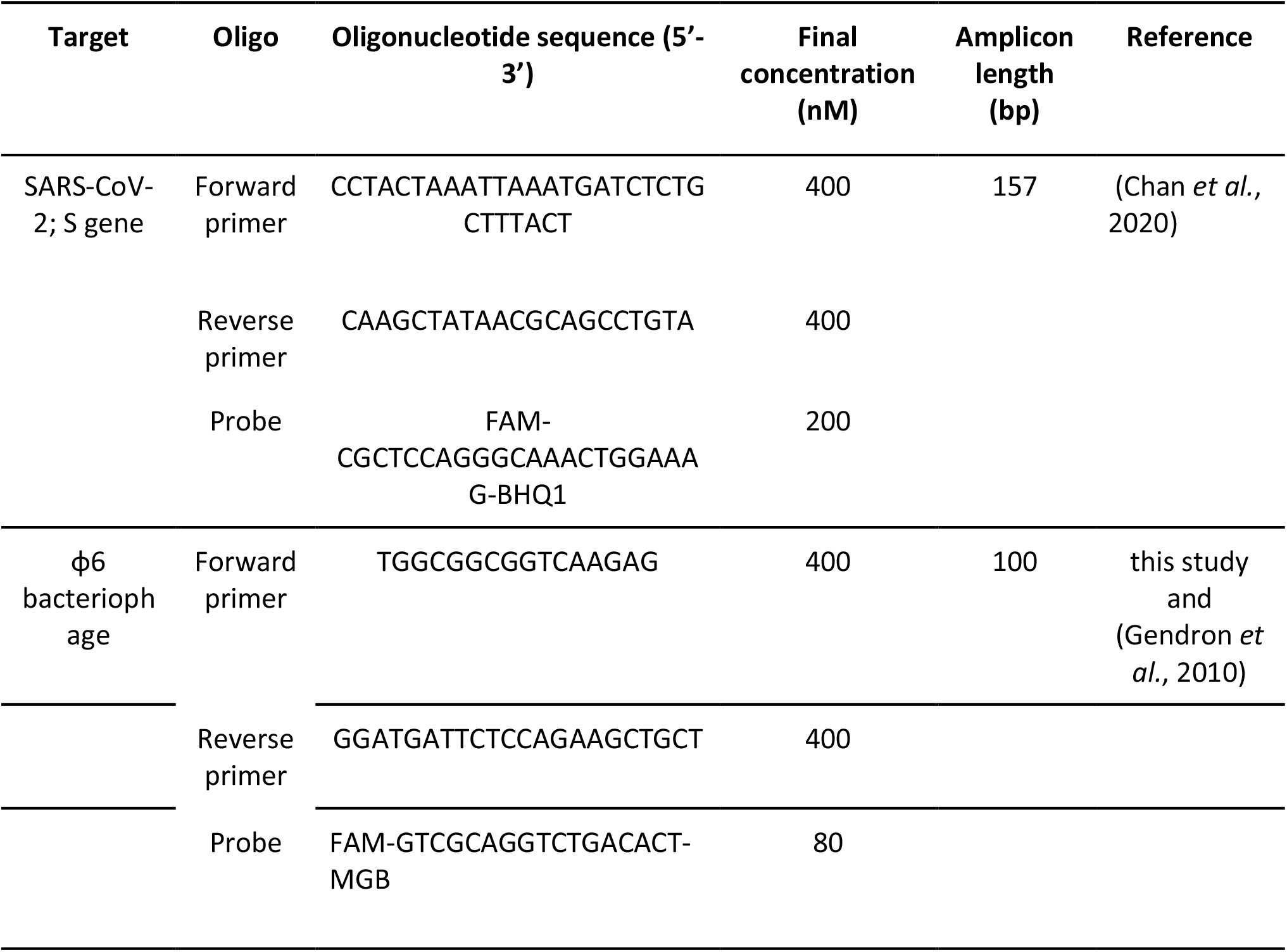
RT-qPCR primers and probe used for the detection of SARS-CoV-2 and Φ6 bacteriophage in environmental samples.

### 2.5 Clinical testing and reporting of COVID-19 positive individuals by the schools

Schools A, B, C, D, and E were offered weekly free COVID-19 clinical testing through HDT (*Healthy Davis Together - Working to prevent COVID-19 in Davis*, 2020; Hubler, 2021). Additional on-site clinical testing was offered at Schools D and E. Positive COVID-19 cases and changes in on-site clinical testing participation were provided to us by administrators at each school or from the school district.

## 3 Results and Discussion

### 3.1 Validation of environmental detection of SARS-CoV-2 RNA after known exposures

Detection of SARS-CoV-2 RNA on environmental surfaces using RT-qPCR has been demonstrated for a wide range of contexts and surface types (Zhou *et al*., 2020; Abrahão *et al*., 2021; Coil *et al*., 2021; Horve *et al*., 2021). We validated our sampling and analytical protocol by sampling in two locations (a private residence and a school classroom) where at least one COVID-19 positive individual was known to have been present within the past week. First, we sampled surfaces at a house where an asymptomatic person who later tested positive for COVID-19 was present for 2 hours. Of the ten surface swab samples collected four days after the exposure, the underside of the chair where the COVID-19 positive person had sat and the floor underneath this space were positive for the virus (Table S2), with Ct values of 37.2 and 37.6, respectively. Our results confirmed that SARS-CoV-2 RNA could be detected on surfaces a few days after the exposure, including surfaces that are not frequently touched (e.g., the floor). High-frequency touched surfaces in workplace environments have tested positive for the SARS-CoV-2 virus days after the detection of a positive individual (Marshall *et al*., 2020). Positive samples have also been collected from no-touch surfaces up to 27 days after an individual was diagnosed with COVID-19 (Dumont-Leblond *et al*., 2021).

Second, we sampled a classroom where two students tested positive after attending school. We conducted two sampling episodes, two and six days after the last day of student attendance. In both sampling episodes, the undersides of the chair, desk and tool box of one of the COVID-19 positive students were positive for SARS-CoV-2 (Table S2), with Ct values of 35.6, 36.8, and 37.6 during the first episode, and 36.2, 36.5, and 36.8 during the second episode, respectively. A portable HEPA filter that was operating in the room during and prior to the exposure was dismantled, swabbed, and tested for SARS-CoV-2. Three samples were collected from each of the outer grill cover and pre-filter mesh, and four samples from the H13 HEPA filter. One, three and two of those samples, respectively, were positive for SARS-CoV-2 (Table S2), with Ct values ranging from 36.1 to 38.7. Twenty other samples collected in the classroom during the two sampling episodes tested negative for SARS-CoV-2 (Table S2).

These preliminary results provided further evidence for the stability of SARS-CoV-2 RNA in the environment at least six days after deposition and validated the potential use of portable HEPA filters for environmental monitoring of COVID-19. Environmental monitoring for the presence of SARS-CoV-2 RNA through air sampling has been demonstrated in clinical and transportation settings using diverse air samplers (Barbieri *et al*., 2021; Moreno *et al*., 2021), and by swabbing or vacuuming HVAC systems (Maestre *et al*., 2021; Moreno *et al*., 2021). However, acquiring and deploying air samplers is challenging due to high costs and noise levels of these instruments. HVAC systems can be difficult to access and interpreting results can be challenging due to shared airflow among different rooms. Deploying HEPA filters is a lower-cost, more accessible, quieter, and better-targeted alternative to using air samplers or HVAC systems for environmental monitoring. Deploying HEPA filters for environmental monitoring has the added benefit of reducing risk of airborne SARS-CoV-2 transmission (Curtius, Granzin and Schrod, 2021; Rodríguez *et al*., 2021).

### 3.2 Environmental monitoring for SARS-CoV-2 in K-6 and K-8 schools using floor swab samples

We partnered with five schools to implement surface-based SARS-CoV-2 environmental monitoring. In School A, eighteen rooms were monitored weekly from January 28 to August 12, 2021 through floor swabbing. SARS-CoV-2 positive environmental samples were intermittently detected only in Room 7 on March 11, April 22, and May 13, 2021 with negative results in between these dates on April 1, 8, 15, and 29, 2021 (Figure 1A). Two of 18 floor samples collected on May 6, 2021 in School A were also positive, but the tube labels were unidentifiable (excluded from Figure 1A). A fresh set of samples collected from the 18 rooms on May 10 were negative. No clinical positive cases were reported from School A classrooms when surface samples were positive. However, an individual from Room 7 was confirmed to be positive for COVID-19 in early February, 2021 (personal communication with school administrator), close to a month before the first positive environmental detection on March 11, 2021. Relic SARS-CoV-2 RNA, i.e., RNA in the environment from degraded and non-infectious virus that has little significance to public health, has been detected from a few weeks after the recovery or termination of quarantine for patients (Fernández-de-Mera *et al*., 2021; Liu *et al*., 2021), and up to two months after symptom onset in a single household with two isolated patients (Maestre *et al*., 2021). As all identifiable positive environmental samples in School A originated from a single room and showed intermittent positive results throughout a period of more than two months, it is possible that relic RNA shed weeks before the initial detection led to intermittent positive tests during our pilot study. Air filter sampling from single HEPA units in all rooms in School A started on May 20, 2021 in parallel to the floor sampling. All HEPA filter and floor samples collected between May 20th and August 12, 2021 were negative for SARS-CoV-2.

**Figure 1.**
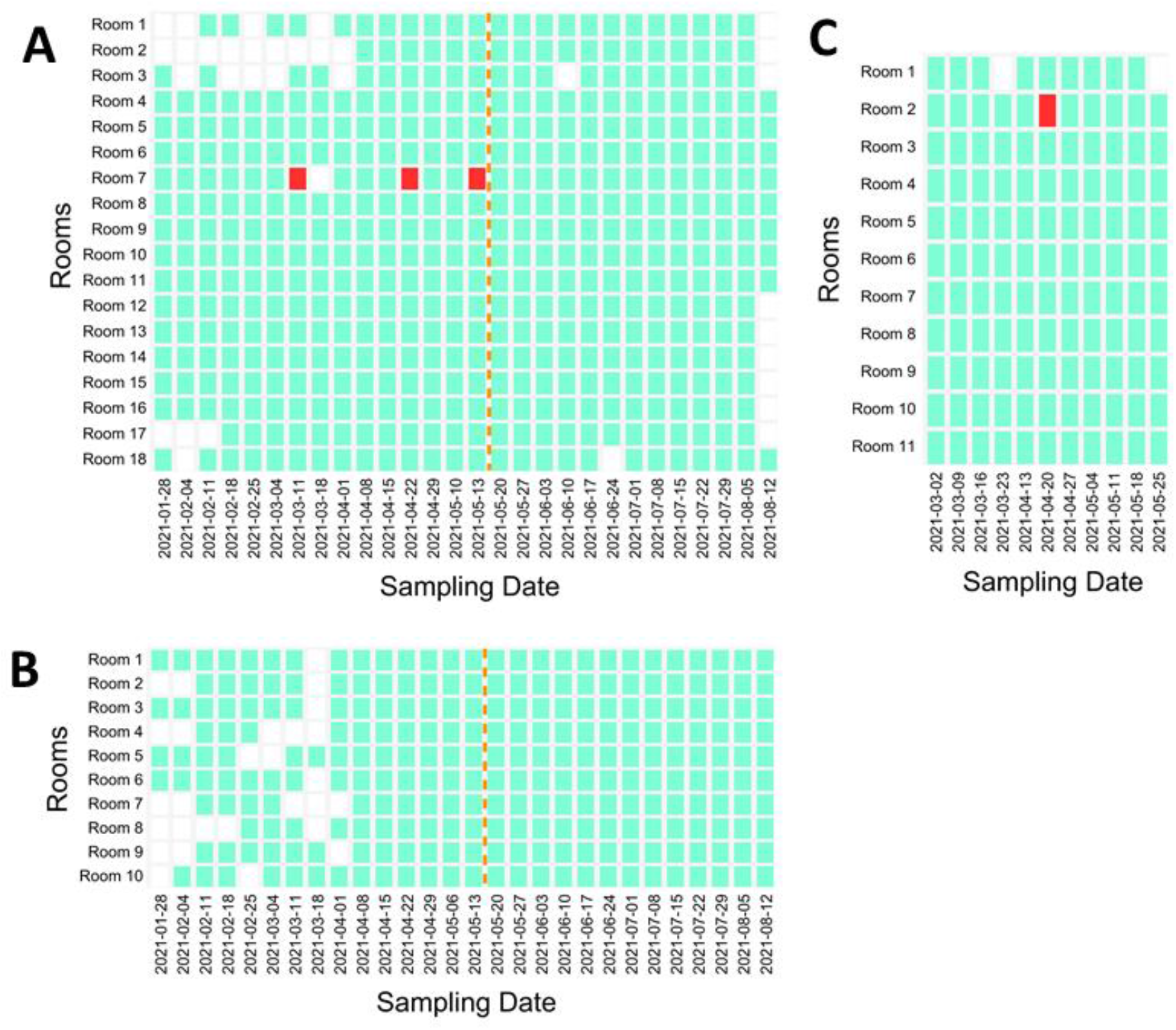
Positive and negative rooms for SARS-CoV-2 based on floor samples collected in Schools (**A**) A, (**B**) B, and (**C**) C throughout the pilot environmental monitoring study. Episodes with a positive floor sample are marked in red, negative episodes in green, and episodes where no sample was collected are in white. Air filter sampling in Schools A and B started on May 5, 2021 and is denoted by the orange line. No positive air filter samples were detected.

In School B, ten rooms were monitored from January 28 to August 12, 2021 through floor swabbing. No positive floor samples for SARS-CoV-2 were detected (Figure 1B). Air filter sampling of single HEPA units in all rooms except Rooms 2, 4, and 7 began on May 20 and continued until August 12, 2021 with no positive samples detected.

In School C, eleven rooms were monitored weekly from March 2 to May 25, 2021 through floor swabbing. Preliminary sampling was conducted on February 23, 2021 when diverse surfaces were sampled in the same eleven rooms, and all of these samples tested negative. The only positive sample for SARS-CoV-2 throughout the weekly sampling was collected in Room 2 on April 20, 2021 (Figure 1B). Unlike our results in School A, no repeated positives in the same room were observed. All teachers, staff and students associated with the positive room were tested for COVID-19 after the positive floor swab, but no clinical cases were found. A family member of a Room 2 occupant did test positive following the environmental detection (personal communication with school administrator). This positive test for a family member raises an important possibility that needs to be considered: SARS-CoV-2 RNA could be shed by an “outsider” (i.e., not someone in the school) and then brought into the sampled environment by someone who is not actively shedding themselves. The mechanical transfer of SARS-CoV-2 RNA on fomites has not been rigorously tested; however, viral RNA has been detected on personal items like clothes, towels, bedding, mobile phones, and shoe soles (Jiang *et al*., 2020; Santarpia *et al*., 2020; Goodwin *et al*., 2021; Redmond *et al*., 2021). Transfer of outsider SARS-CoV-2 genetic material to sampled rooms is thus an important possibility that should be considered in the context of environmental monitoring.

### 3.3 Floor and HEPA filter swab sampling for the detection of SARS-CoV-2

We established a different environmental monitoring strategy in Schools D and E, informed by the results from the HEPA filter sampling after a known exposure test detailed in section 3.1. From the first day of sampling, we collected one floor and two HEPA filter samples (one sample from each of two HEPA filtration units) per room. In School D, preliminary testing episodes were conducted in Rooms 11 and 20 on March 10, 16 and 30, 2021. All HEPA filter and floor samples collected during these episodes were negative for SARS-CoV-2. The weekly sampling in School D covered 25 rooms and ran from April 13 to June 8, 2021. Ten HEPA filter samples and one floor sample collected during this period tested positive (Figure 2A). All positive detections were from a single filter or floor sample at a time; that is, no more than one sample per room ever tested positive for SARS-CoV-2 during the same sampling episode. Positive air and surface samples in schools have been previously reported (Cordery *et al*., 2021; Crowe *et al*., 2021). Air sampling through the use of air filter samplers, electrostatic precipitators and HVAC filters has been demonstrated (Barbieri *et al*., 2021; Crowe *et al*., 2021; Maestre *et al*., 2021; Moreno *et al*., 2021). To the best of our knowledge, this is the first study to report portable HEPA filter sampling as a strategy for detecting environmental SARS-CoV-2 RNA. Limited participation in on-site clinical testing was observed after the reporting of positive environmental results on most episodes; therefore, no direct links were established between environmental and clinical results. Participation in testing varied between schools and no data is available for schools A, B, and C. Testing in School D varied week to week from 33% to 88% with similar but slightly lower numbers at School E.

**Figure 2.**
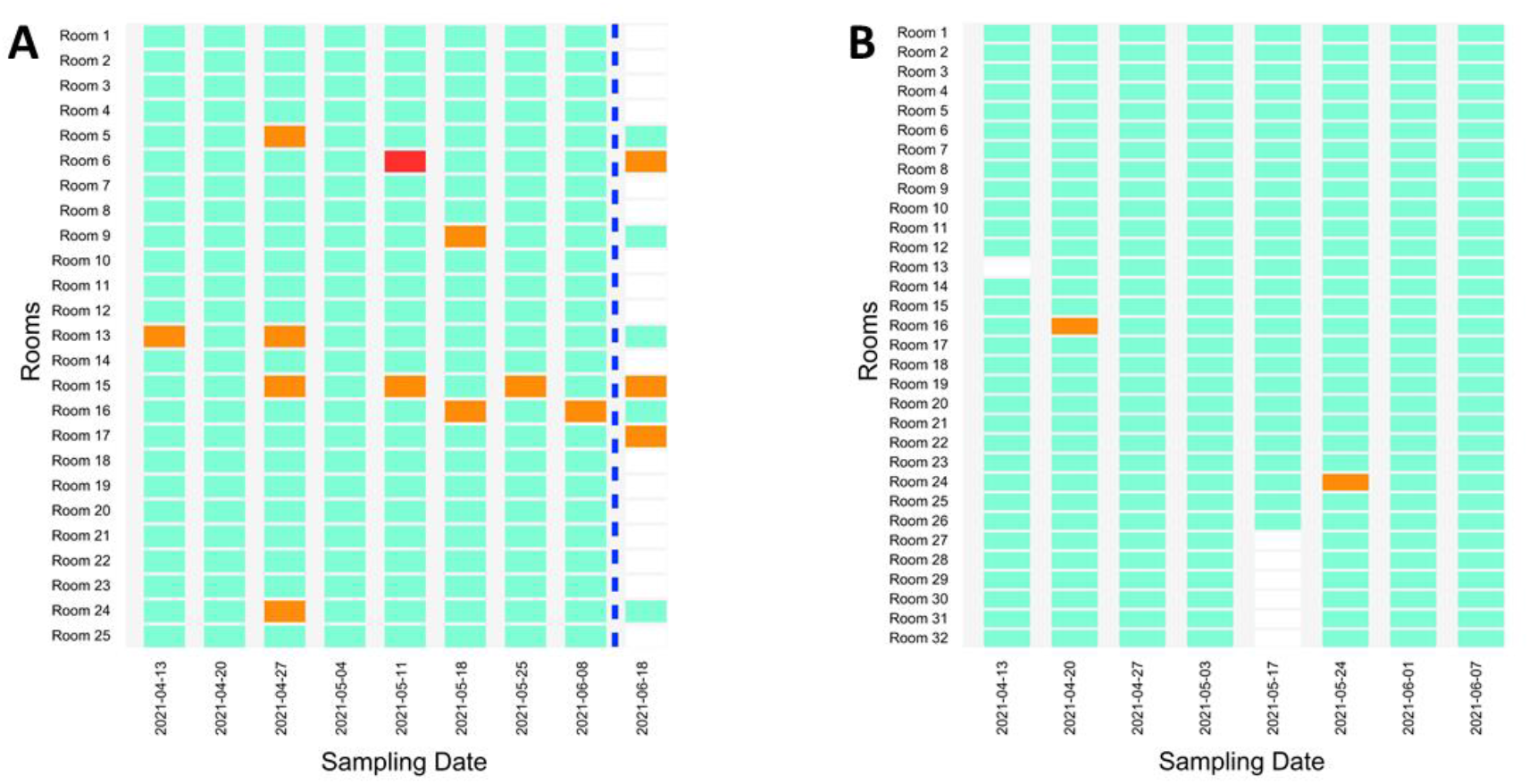
Positive and negative rooms for SARS-CoV-2 based on floor and HEPA filter samples collected in (**A**) School D and (**B**) School E throughout the pilot environmental monitoring study. Testing dates with a positive floor sample are in red, episodes with a positive air filter sample are in orange, negative episodes are in green, and episodes where no samples were collected are in white. No more than one sample tested positive in a room at any given time. A concluding sampling episode was conducted in School D on June 18, 2021 after school sessions ended to gather further information on the persistence of environmental SARS-CoV-2 RNA in previously positive classrooms and is indicated by the blue line. The previously negative Room 17 was included in the concluding sampling episode as a negative control. Six samples collected in School E on May 17, 2021 were impossible to link to specific rooms because the tube labels were compromised; however, the samples were collected from Rooms 27-32 and all tested negative for SARS-CoV-2 (not included in the figure).

A concluding sampling episode was conducted in School D on June 18, 2021 eight days after the last day of the school year. The goal of this sampling episode was to gather further information on the persistence of environmental SARS-CoV-2 RNA in previously positive classrooms. Rooms 5, 6, 9, 13, 15, 16, 17 and 24 were sampled. Each of these rooms except for Room 17 (selected as a negative control) yielded a positive detection of SARS-CoV-2 at some point during the weekly sampling. During this episode, samples from a door jamb, teacher desk and center surface (either a desk, chair, podium, or projector located in the center of the classroom) were collected from each room alongside the two filter samples and single floor sample. One filter sample from Room 6 and one filter sample from Room 15 tested positive once more (Figure 2A). Unexpectedly, a filter sample from Room 17 (the negative control) tested positive as well. These results further confirmed the challenges of interpreting positive results in previously positive environments, since positive results could be caused by the resuspension and capture of relic RNA on air filters.

In school D, we implemented HEPA filter replacement after a positive detection to mitigate the impact of relic RNA contamination on future samples and avoid the repeated positives issue observed in School A. The strategy was not successful as repeated SARS-CoV-2 positives were observed in three rooms after new filters were installed during the April 13 to June 8, 2021 sampling period. Rooms 6, 13, 15, and 16 yielded two or four repeated intermittent positives during the pilot study, including the concluding sampling episode (Figure 2A). The longest period of intermittent repeated positives was in Room 15, covering 52 days. Even with the change in filters after a positive environmental detection, capturing relic RNA through HEPA filter sampling remained a possibility. The intermittent and repeated positives could have been a result of the resuspension of dust particles containing SARS-CoV-2 genetic material. SARS-CoV-2 RNA has been detected in floor and HVAC dust up to two months after patient symptom onset (Maestre *et al*., 2021), which could explain our results in the absence of clinical confirmation due to the low testing participation.

In School E, we conducted preliminary testing episodes on March 10, 16, and 30, 2021 in a room that was not part of the weekly sampling campaign. The air filter and floor samples collected during the preliminary testing were negative for SARS-CoV-2. Weekly sampling covering 32 rooms was conducted from April 13 to June 7, 2021. As in School D, we collected two HEPA filter samples and one floor sample from each School E classroom during each sampling episode. A HEPA filter replacement strategy was also implemented in School E. Of all the samples collected during the campaign, only two—a single filter sample each from Rooms 16 and 24—tested positive for SARS-CoV-2 (Figure 2B). No repeated positives were observed. This cannot be attributed to the HEPA filter replacement strategy as results from School D show that relic RNA capture on new filters is possible. Site-specific conditions may contribute to the resuspension of dust particles, including room ventilation (X. Wang *et al*., 2000), which could explain the fact that intermittent positives were observed in some classrooms but not others. Increased on-site clinical testing participation was observed in School E after the positive environmental results but no positive individuals were identified.

### 3.3 High Ct values in floor and air filter swab samples

qPCR Ct values are inversely proportional to the concentrations of the target genes in the samples tested. Observed Ct values across all filter and floor samples collected for this study ranged from 36.07–38.01 (Figure 3A and 3B), which is close to a commonly used threshold for considering a sample positive (Ct<40) (Dumont-Leblond *et al*., 2021; Harvey *et al*., 2021). The high Cts obtained were likely due to low amounts of virus collected through the environmental sampling methods. These Ct values are in the range of what has been observed for SARS-CoV-2 surface sampling, with values of >30 in a clinical environment with COVID-19 patient and non-patient care areas (Zhou *et al*., 2020), a 35 and 34–36.5 median and interquartile range, respectively, in quarantine environments (Liu *et al*., 2021), 34-44 in schools (Cordery *et al*., 2021), 34-38 in workplace sites (Marshall *et al*., 2020) and 29.0-38.1 in public locations such as public squares and bus terminals (Abrahão *et al*., 2021). Similarly, Ct values in the 36 to 39 range have resulted from air sampling with glass fiber filters in a clinical setting (Barbieri *et al*., 2021).

**Figure 3.**
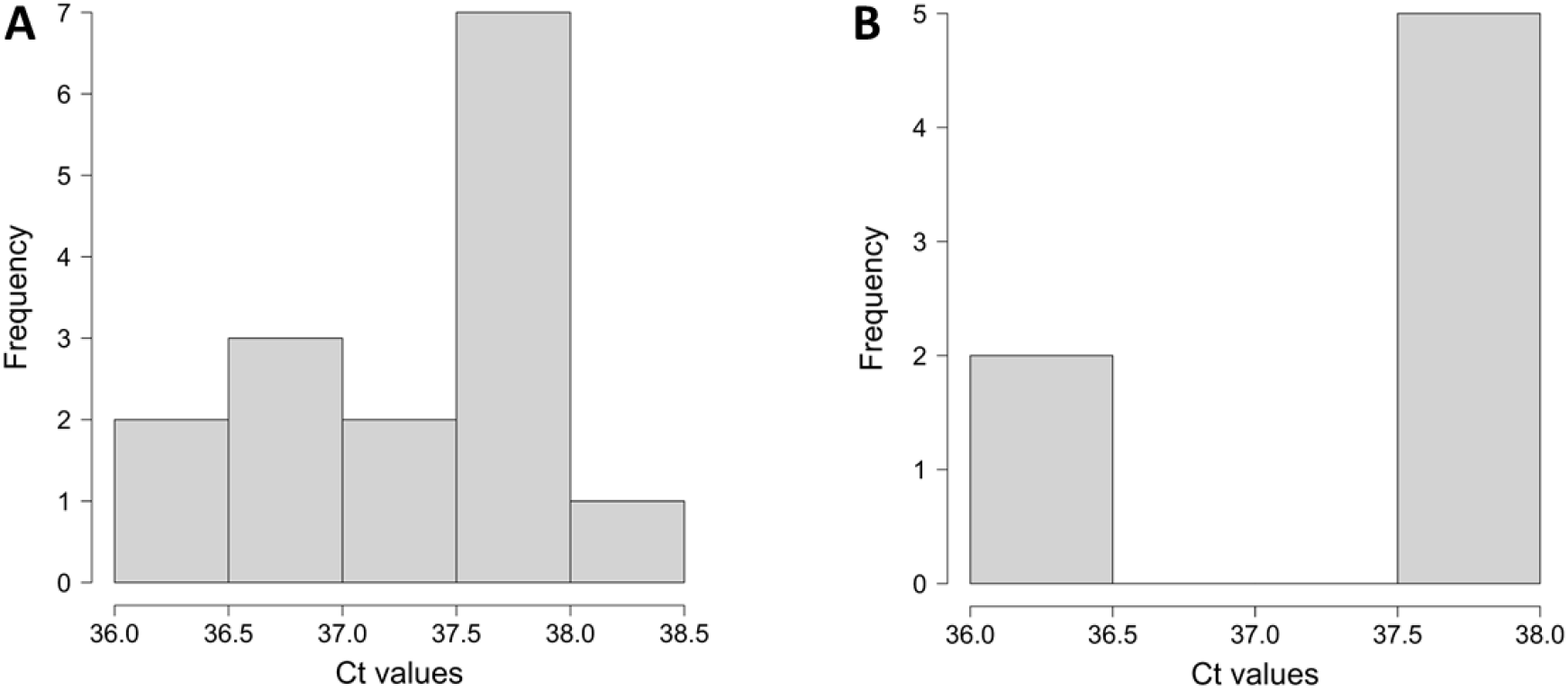
Frequency distributions of cycle threshold (Ct) values for positive (**A**) HEPA filter and (**B**) floor samples obtained from Schools A, B, C, and D. All positive samples were processed using the MagMAX automated extraction protocol and tested using a SARS-CoV-2 S gene RT-qPCR assay.

In the present study, air filter Ct values were more evenly distributed across the 36.37-38.01 Ct range (Figure 3A) than floor samples. Positive floor samples had Ct values that ranged between 36.07-36.35 and between 37.66-37.85 (Figure 3B). For both air filter and floor positive samples, 12 out of the 22 Ct scores were in the 37.5-38.0 range. The similar Ct results in air filter and floor samples suggest comparable amounts of virus deposited on the two surface types as the same sample collection method was used for both strategies. Infectious virus has not been recovered from environmental samples with Ct values above 30 (Zhou *et al*., 2020; Krambrich *et al*., 2021), and although possible, further testing is required to assess the state of virus viability on diverse surfaces. The high Ct values obtained in our study also pose a challenge for differentiating fresh from relic SARS-CoV-2 RNA. It is possible that all the positives detected were from relic genetic material as no infected individuals were found through clinical testing, although with the limited participation observed.

### 3.4 Surface and air filter sampling as strategies to inform the management of the COVID-19 pandemic

Even though no clinical positive cases of COVID-19 were identified following the detection of environmental positives in Schools D and E, clear differences were observed in the environmental positivity rates of the two schools. For School D, the mean environmental positivity rate was 5.5% positive rooms per sampling episode (considering filter and floor samples together, and excluding the concluding sampling session). This positivity rate is seven times higher than the 0.78% positivity rate observed for School E. Self-reported positive case data from Schools D and E for the 2021 school year was provided to us by the school district. No positives were reported during the environmental sampling period (April 13 to June 8, 20201) in either of the two schools. However, two positive cases were reported in School D in early February, during limited in-person instruction. No in-person instruction cases were reported in School E throughout the school year. The larger number of prior clinical positive cases and subsequent higher environmental positivity rates in School D highlight the potential link between clinical and environmental results, although they also accentuate the possible role that relic RNA can have on the observed results. Relationships between environmental surface positivity rates and clinical cases in larger populations have been established (Harvey *et al*., 2021), showing the potential for environmental monitoring regardless of clinical testing participation. In the present study, the number of positive rooms in schools D and E, eight and two respectively, was higher than the number of self-reported cases (including prior cases). Incomplete testing penetration and relying on self-reporting are likely to miss positive cases. In these situations, the environmental positivity rate may be a useful statistic to inform pandemic management strategies in the absence of a better metric.

The two strategies utilized in this study for SARS-CoV-2 monitoring in the environment, namely floor and HEPA filter sampling, yielded different positivity rates. There were 15 identifiable air filter positives throughout the sampling period in Schools D and E (including the final sampling episode in School D) but only a single positive floor sample. However, two HEPA filter samples were collected from each room, compared to one floor swab. Taking into account the larger sample size, the HEPA filter positivity rates in Schools D and E were 3.02% and 0.41%, respectively, while the floor positivity rates were 0.48% and 0%. These results suggest that HEPA filter sampling is more sensitive than floor sampling. This can be explained because air filters collect and concentrate particles that can contain SARS-CoV-2—including aerosols, droplets, and dust—on a relatively small surface area. These types of particles also deposit on floors, although collecting one sample from larger areas has a lower probability of picking up deposited SARS-CoV-2 RNA. This surface sampling challenge has been previously reported in a high-frequency touched surfaces study (Marshall *et al*., 2020). Concentration is desired for environmental samples and has been shown to allow a higher sensitivity for the detection of SARS-CoV-2 in samples such as wastewater (Jafferali *et al*., 2021; LaTurner *et al*., 2021). Finally, floors are cleaned more frequently than HEPA filters are replaced—although, as previously noted, we observed repeated detection of SARS-CoV-2 in School D classrooms even after filters were replaced.

There are clear challenges with using surface and filter sampling for monitoring the presence of SARS-CoV-2. Many of these challenges, including near-limit-of-detection virus concentrations and the impacts of site-specific conditions, have also been reported for other types of environmental monitoring, such as wastewater surveillance (Hart and Halden, 2020). However, the biggest challenge we encountered in our study was the differentiation between freshly shed, relic or outsider SARS-CoV-2. Based on our results, monitoring surfaces and air filters for SARS-CoV-2 can only be complementary to clinical testing if the goal is to find the source of the virus detected. However, when implementing this type of environmental monitoring, it has to be clear that individuals may test negative as the environmental detection could have been due to relic or outsider RNA. Clinical testing as a response to relic or outsider environmental RNA can be useful as it provides certainty that all individuals in the room are negative for the virus. SARS-CoV-2 environmental surface monitoring should also be complementary to clinical testing, if available, because viral shedding can be too low to be detected environmentally; issues that have been described previously (Marshall *et al*., 2020; Ryu *et al*., 2020). If full clinical testing participation is not possible nor available, SARS-CoV-2 environmental surface monitoring is limited to detecting, but not differentiating between freshly shed, relic, or outsider RNA. This strategy can still provide useful information for the pandemic management by setting baseline environmental positivity rates, tracking rate changes through time and responding to those changes with mitigation measures.

## 4 Conclusions

Portable HEPA filter and floor sampling are environmental monitoring tools that can successfully detect SARS-CoV-2 RNA. HEPA filter sampling appears to be a more efficient tool compared to floor swabs. In schools or other settings where access to or participation in clinical testing programs is limited, HEPA filter testing could be a useful strategy to inform pandemic response. However, environmental monitoring of SARS-CoV-2 through surface sampling (including HEPA filters) poses the challenge of differentiating amongst fresh, relic, and outsider viral RNA, especially for high-Ct results. Further research is needed to establish Ct thresholds for HEPA filter monitoring that indicate nearby active infections and elevated exposure risks. New technologies and testing protocols that differentiate fresh from aged viral RNA would also do much to increase the utility of SARS-CoV-2 environmental monitoring in schools.

## Data Availability

All data produced in the present work are contained in the manuscript

## Acknowledgements

The authors would like to thank the many teachers, administrators, staff, parents and others associated with these schools for facilitation of sample collection. We also thank the The Biology and Built Environment Center at the University of Oregon for supplying expertise and reagents at the start of this project. Funding was provided by Healthy Yolo Together/Healthy Davis Together, a partnership between the University of California, Davis, the city of Davis, and Yolo county. Members of the Bischel lab provided valuable laboratory support and feedback on the manuscript.

## Supplementary information

### S1 Supplementary Methods

#### S1.1 Validation sampling for the environmental detection of SARS-CoV-2 on surfaces and HEPA filtration units

We validated the detection of SARS-CoV-2 through swabbing surfaces and HEPA air filter units in two locations in Northern California after known exposures. The first location was a house where a visitor stayed for a period of 2 hours, mostly wearing a mask with the exception of ∼20 minutes to have a meal. The visitor received a positive COVID-19 test result one day after the visit. We obtained swab samples from different surfaces in the dining, living and bath rooms four days after the exposure (Table S2). At the second location, which was a school classroom, two students tested positive for COVID-19 before the cohort was quarantined. We sampled surfaces two and six days after the students were on campus (Table S2). A portable air purifier equipped with an H13 HEPA filter (MA-40, Medify Air, USA) that was active in the room when the positive individuals were present was dismantled and parts of it were sampled three days after. Three samples each were collected from the outer unit grill cover and the pre-filter mesh, and four samples were collected from the H13 HEPA filter surface (Table S2).

#### S1.2 SARS-CoV-2 floor and HEPA filter sampling instructional videos

Floor sampling instructional video can be found here: https://youtu.be/HuOuzR9Rpg8

HEPA filter sampling instructional video can be found here: https://youtu.be/MzV8tDMKsZc

#### S1.3 Detailed RNA extraction protocols

We used two methods to extract the SARS-CoV-2 RNA from surface and HEPA filter samples, one manual and one automated. Section 2.4 details the periods of time when each method was utilized. For the manual extractions with the PureLink Viral RNA/DNA kit (Thermo Fisher Scientific, USA), sample volumes of 200 μl were mixed with 25 μl of Proteinase K and 200 μl of Lysis Buffer. The resulting solutions were vortexed for 15 seconds and incubated at 56 °C for 15 minutes. 250 μl of 100% ethanol was added, vortexed for 15 seconds and incubated at room temperature for 5 minutes. The solutions were centrifuged for 20 seconds at 10,000 x g to remove foam. The lysates (∼675 μl) were transferred to Viral Spin Columns in collection tubes and centrifuged for 1 minute at 6,800 x g. The Spin Columns were placed in new Wash Tubes. 500 μl of Wash Buffer (WII with ethanol) was added to the Spin Columns and centrifuged for 1 minute at 6,800 x g. The flow through was discarded, and the 500 μl Wash Buffer (WII with ethanol) wash and centrifugation were repeated. The Spin Columns were placed in clean Wash Tubes and centrifuged for 1 minute at maximum speed (∼20,000 x g) to remove residual Wash Buffer. The Spin Columns were placed in sterile microcentrifuge tubes and 50 μl of sterile RNase-free water was used to elute the RNA extracts. The samples were incubated at room temperature for 1 minute and centrifuged at maximum speed to elute the RNA. The extracts were stored in a -80 °C freezer.

The MagMAX Microbiome Ultra Nucleic Acid Isolation Kit (Applied Biosystems, USA) and the KingFisher Flex purification system (Thermo Fisher Scientific, USA) were used for automated RNA extractions. Sample volumes of 200 μl were transferred to 96-Deep Well Kingfisher plates. A solution that contained 250 μl of MagMAX Viral/Pathogen Binding Solution and 10 μl of MagMAX DNA/RNA Binding Beads per sample was made and 260 μl of the solution was added to each well of the 96-Deep Well plates containing samples. Wash 1 and Wash 2 Kingfisher plates were prepared with 1 ml of MagMAX Viral/Pathogen Wash Solution per well. Wash 3 and Wash 4 plates were prepared with 1 ml of 80% ethanol per well. Elution plates were prepared with 100 μl of MagMAX Viral/Pathogen Elution Buffer in each well. The plates were loaded into the Kingfisher and the MagMAX_Microbiome_Stool_Flex.bdz extraction protocol was run. The extracted samples were transferred to microcentrifuge tubes for storage at -80 °C.

#### S1.4 RT-qPCR standard curves

Standard curves were created for the spike glycoprotein (S) and Φ6 bacteriophage assays. Known copy numbers of SARS-CoV-2 RNA obtained from clarified viral supernatants of heat inactivated virus donated by the University of Oregon were used for the SARS-CoV-2 RNA standard curve. Four replicates of ten-fold serial dilutions of viral RNA in the range of 3.2×10^5^ to 3.2 copies per μl were used for generating the standard curve (Table S1). The primers and probes detailed in Table 1 were used following the thermal cycling conditions as in section 2.4. The lower limit of detection was determined as the lowest concentration at which all four replicates amplified.

### S2 Supplementary tables

**Table S1.** RT-qPCR standard curve for SARS-CoV-2 using viral RNA.

| Target | Standard curve | R<sup>2</sup> | Efficiency<sup>a</sup> | Limit of detection |
| --- | --- | --- | --- | --- |
| SARS-CoV-2<br>S gene | y= -3.218x + 39.052 | 0.999 | 104.53% | 3.2 copies/μl of<br>extract |
| φ6<br>bacteriophage | y=-2.961x + 38.822 | 0.998 | 117.65% | 11.6 copies/μl of<br>extract |

<sup>a</sup>A PCR efficiency = $10^{-1/\text{slope}} - 1$

**Table S2.**
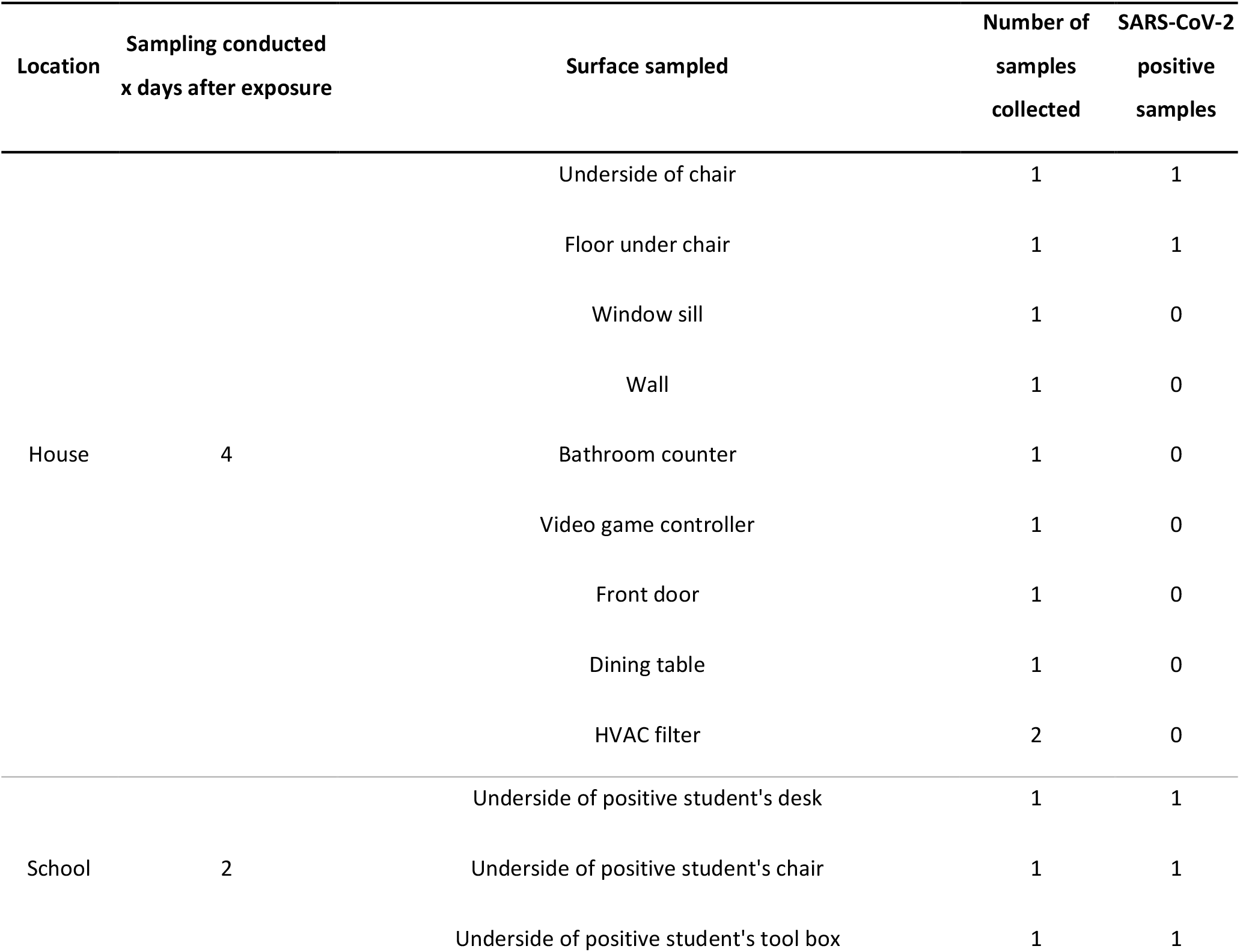
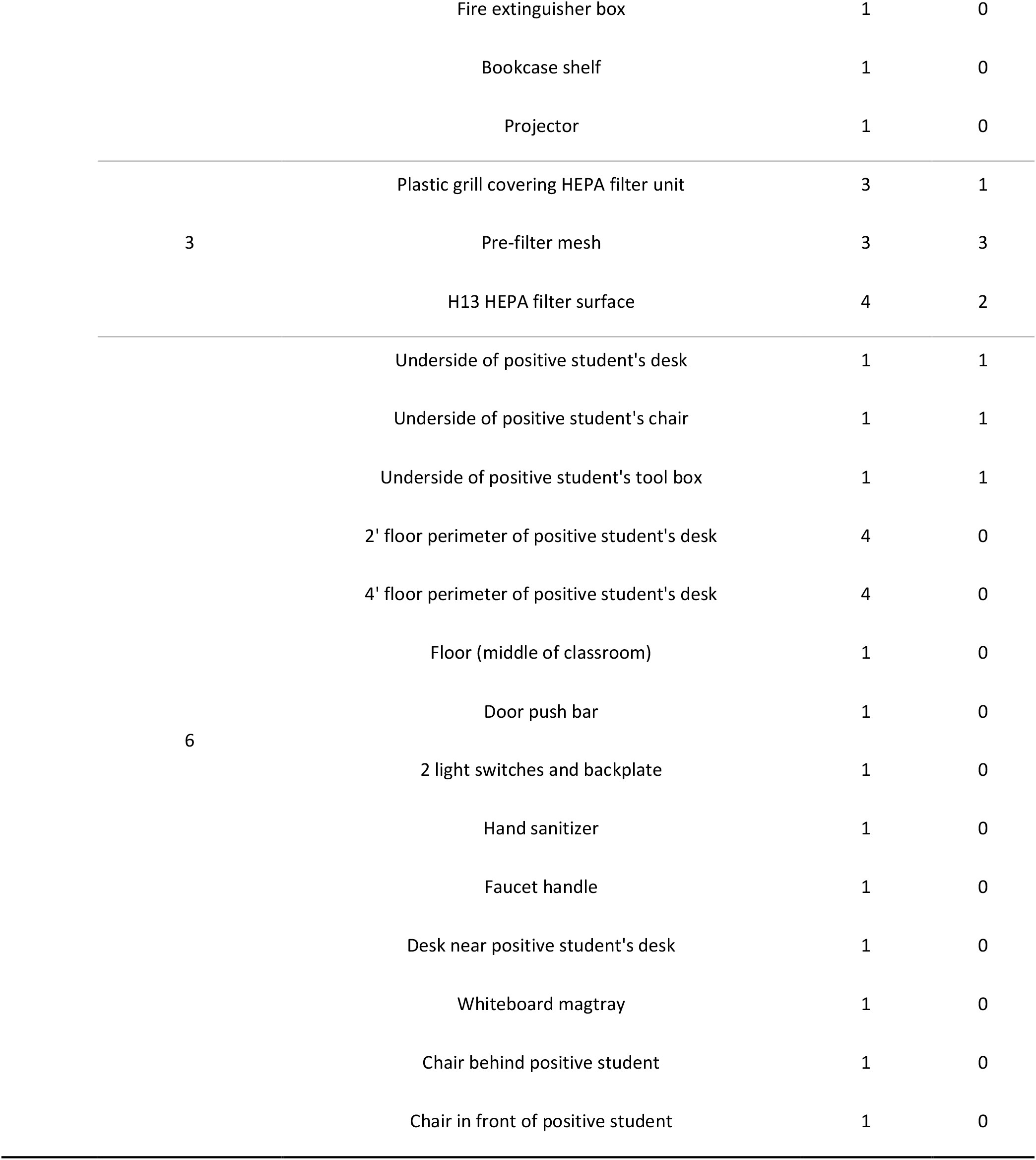
Samples tested during the validation experiments for the detection of SARS-CoV-2 on surfaces and HEPA air purification units after known exposures.

